# Assessing Knowledge, Attitudes, and Practices towards Causal Directed Acyclic Graphs: a qualitative research project

**DOI:** 10.1101/2020.01.17.20017939

**Authors:** Ruby Barnard-Mayers, Ellen Childs, Laura Corlin, Ellen C Caniglia, Matthew P Fox, John P. Donnelly, Eleanor J Murray

**Author notes:** Corresponding Author: Ruby Barnard-Mayers, Department of Epidemiology, Boston University, Boston, MA, 02118. **Declarations** *Ethics Approval* The survey was exempted by the Boston University School of Public Health Institutional Review Board. The surveys were completely anonymous and no IP addresses were collected. Funding* EJM and RBM were partly funded by the Eunice Kennedy Shriver National Institute of Child Health and Human Development (NICHD) R21HD098733. Availability of data and material* The full survey used to obtain these results is included in the supplementary material. **Author Contributions** In terms of author contributions for this paper; RBM & EJM conceived of the idea; RBM, EJM & EC designed the survey; and all authors approved the survey instrument & contributed to the manuscript writing.

## Abstract

**Background:** Causal graphs provide a key tool for optimizing the validity of causal effect estimates. Although a large literature exists on the mathematical theory underlying the use of causal graphs, less literature exists to aid applied researchers in understanding how best to develop and use causal graphs in their research projects.

**Methods:** We sought to understand why researchers do or do not regularly use DAGs by surveying practicing epidemiologists and medical researchers on their knowledge, level of interest, attitudes, and practices towards the use of causal graphs in applied epidemiology and health research. We used Twitter and the Society for Epidemiologic Research to disseminate the survey.

**Results:** Overall, a majority of participants reported being comfortable with using causal graphs and reported using them ‘sometimes’, ‘often’, or ‘always’ in their research. Having received training appeared to improve comprehension of the assumptions displayed in causal graphs. Many of the respondents who did not use causal graphs reported lack of knowledge as a barrier to using DAGs in their research.

**Conclusion:** Causal graphs are of interest to epidemiologists and medical researchers, but there are several barriers to their uptake. Additional training and clearer guidance are needed. In addition, methodological developments regarding visualization of effect measure modification and interaction on causal graphs is needed.

## Introduction

Causal inference is a growing field in epidemiologic research. To investigate a causal relationship, researchers must make several assumptions regarding the relationship between covariates, the exposure, the outcome, and the population of interest. Although causal graphs do not change the required assumptions for epidemiologic studies, they are a useful tool for improving the clarity of these assumptions, and simplify the process of questioning the validity of assumptions, and designing appropriate data collection and analytic plans in light of them [1,2]. “*Causal graph*” is an umbrella term for visualizations that explain relationships between variables and provide a theoretical framework on which to base an etiologic study. Directed acyclic graphs (DAGs), single world intervention graphs (SWIGs), decision trees, and finest fully randomized causally interpretable structured tree graphs (FFRCISTGs) are types of causal graphs. A detailed explanation of these types of causal graphs is outside the scope of this paper. Interested readers should refer to *Causal Inference: What If* [3] for a comprehensive description of causal graphs.

Although the mathematical foundations of causal graphs have been well-specified[1,3,4], they have not been widely adopted in epidemiology and medical research [2]. We believe the reason for this implementation lag is a paucity of available tools for the application of causal graphs in the study design and analysis phases of applied research studies, although excellent methodological resources exist [5]. In anecdotal conversations with epidemiologists and other medical researchers, barriers to utilizing these models included uncertainty about which variables within a causal model to include in a statistical model, how to model complex or composite variables, and when to consider multiple time points or temporal ordering, as well as a fear of inviting criticism if the graph is implemented in a way others perceive as incorrect. While there are general mathematical axioms that provide some guidance on these questions, there are few resources that address them on a practical level.

The aim of this project was to characterize the knowledge, attitudes, and practices of epidemiologists and other medical researchers in relation to the use of causal graphs in their applied research studies, focusing on causal DAGs. We conducted a convenience survey of practicing epidemiologists and researchers in closely related fields (e.g., biostatistics, medicine, health services research). The goals of this survey were: (1) to determine knowledge of the basic principles of creating and interpreting causal graphs; (2) to determine attitudes towards, experiences with, and practices of the use of causal graphs for designing applied research studies in order to identify needs and guidance; and (3) to identify perceived barriers to the use of causal graphs.

## Methods

### Participants

Our target population was researchers who work on applied epidemiologic topics, whether or not they identify as epidemiologists, and who conduct at least some research in English. This is a large target population, and we were not able to enumerate the potential participants. Instead, we used two strategies to identify survey participants. First, we disseminated the request for participation through social media, in particular via Twitter, capitalizing on the large community of epidemiologists and medical researchers who use Twitter for academic conversations under the hashtag #epitwitter. Second, we contacted the Society for Epidemiologic Research (SER), the American Public Health Association, the American College of Epidemiology, and the EpiMonitor newsletter to request our survey be sent out to their members/readers. Of these organizations, only SER agreed to distribute our survey request.

### Survey development

The questions used in this survey were the result of conversations and teaching experiences the authors had when discussing causal graphs. The questions in the knowledge portion of the survey were designed based on the teaching experience of the authors and were created to differentiate those who knew basic DAG theory, including assumptions required to draw and interpret DAGs, and those who did not. Best practices for incorporating effect measure modification into DAGs is currently up for debate, even within this team of authors, and the question on this topic was designed to determine the prevalence of different attitudes within the research community. The questions in parts three (attitudes) and four (practices) were designed to understand how participants felt about using DAGs in their own research and what tools they used or wanted to use in order to apply DAG methodology in their future work. Most questions had an “other” option that allowed participants to type in their own response to questions to ensure that we could capture possible responses that we may have overlooked in our built-in response options (see appendix).

### Survey distribution

The original Twitter link was available for four weeks (August 7^th^, 2019 – September 3^rd^, 2019; see appendix). One week before the survey closed, we posted a reminder that the survey would be closing soon to encourage anyone with an incomplete survey to submit. All survey responses were submitted by the time the survey link closed. Data were analyzed the week following survey closure.

The Society for Epidemiologic Research posted the survey link to Facebook and Twitter on November 20^th^, and the survey accessible from those links was available for six weeks (see appendix). The other three organizations either did not respond to requests to distribute the survey or were unable to approve dissemination requests in the proposed time frame of the study.

### Data collection and analysis

The survey was distributed using the Qualtrics platform and began with a description of the survey and informed consent materials. Participants who consented were asked to complete a section with basic demographic information, including gender, race/ethnicity, country where they work, type of organization (academic, governmental/NGO, industry), and highest level of education. For all questions, participants were able to select the option ‘choose not to respond’ or submit a written answer instead of selecting from the list of options. The survey next asked a series of questions designed to assess training, knowledge, comfort with, and preferences for causal graphs in applied epidemiology. The survey was exempted by the Boston University School of Public Health Institutional Review Board. The surveys were completely anonymous and no IP addresses were collected. The full survey text is provided in the eAppendix.

There were two open-ended survey questions asking participants to expand on their discomfort using DAGs and their attitudes about the usefulness of DAGs in a variety of research stages. We then used a grounded approach to code those responses into groups of similar responses, and systematically applied those groupings across all responses for both qualitative questions, using Microsoft Excel [6–8]. We came into the project with no a priori codes or conceptions of what we would find. One research team member reviewed the responses to each answer and developed codes that reflected types of responses. A second team member reviewed the responses and coded text to validate the groupings. Qualitative survey results were collated and major themes and supporting ideas were identified by study personnel. All results were stratified by source of participants (direct social media versus society dissemination).

## Results

The initial survey push (via the original Twitter link) garnered a total of 400 responses, while the Society for Epidemiologic Research-disseminated survey resulted in 39 new responses. Due to the limited number of responses to the Society for Epidemiologic Research-disseminated survey link, and because we could not verify that there was no overlap in respondents (since we did not collect IP addresses), the remainder of this paper will focus primarily on the results from the first, Twitter-based, version of the survey. Most participants identified as white and there were slightly more female-identifying participants. About a third of participants were students, and of those who were not students most worked in academia. Table 1 shows the demographic and educational breakdown of the participants by survey version. The demographics of those who responded to the SER-disseminated survey generally matched the distribution in the original Twitter-disseminated survey.

**Table 1.** Demographic information of survey respondents, stratified by survey version.

|  | **Twitter Respondents**<br>**(N = 400)** | **Society Respondents**<br>**(N=39)** |
| --- | --- | --- |
| **Current gender** | n=383 | n=36 |
| Woman | 205 (54%) | 21 (58%) |
| Man | 167 (44%) | 15 (42%) |
| Non-binary, gender-fluid, or other | 5 (1%) | 0 (0%) |
| Prefer not to say | 6 (2%) | 0 (0%) |
| **Race / ethnicity [select all that apply]** | n=411 | n=40 |
| Black, including African American & African | 17 (4%) | 1 (3%) |
| Asian, including East Asian and South Asian | 52 (13%) | 6 (15%) |
| Middle Eastern or North African | 11 (3%) | 3 (8%) |
| Indigenous, including Native American, First Nations, Aboriginal, Metis; Pacific Islander or Native Hawaiian | 5 (1%) | 0 (0%) |
| White | 286 (70%) | 26 (65%) |
| Hispanic | 24 (6%) | 3 (8%) |
| Other | 9 (2%) | 1 (3%) |
| Prefer not to say | 7 (2%) | 0 (0%) |
| **Primary Location** |  |  |
| United States | 201 (54%) | 16 (44%) |
| Canada | 24 (6%) | 4 (11%) |
| Other/prefer not to say | 141 (38%) | 16 (45%) |
| Prefer not to say | 9 (2%) | 0 (0%) |
| **Current student status** | n=383 | n=36 |
| Yes | 131 (34%) | 14 (39%) |
| No | 252 (66%) | 22 (61%) |
| **Program of study, current students** | n=128 | n=14 |
| Masters of Science | 9 (9%) | 1 (7%) |
| Masters of Public Health | 8 (8%) | 1 (7%) |
| PhD, ScD, or equivalent | 79 (77%) | 12 (86%) |
| MD; Combined MD-PhD or equivalent | 3 (2%) | 0 (0%) |
| Other or prefer not to say | 3 (2%) | 0 (0%) |
| **Highest level of education, non-students** | n = 245 | n = 22 |
| Master of Science or Master of Arts | 13 (7%) | 3 (14%) |
| Master of Public Health | 13 (7%) | 0 (0%) |
| PhD, ScD, or equivalent | 130 (70%) | 12 (55%) |
| Medical Doctorate; combined MD-PhD or equivalent | 25 (13%) | 6 (27%) |
| Other / prefer not to say | 6 (4%) | 0 (0%) |
| **Field of study (n = 373, 36)** | n=373 | n=36 |
| Epidemiology | 202 (54%) | 22 (61%) |
| Public Health | 43 (12%) | 5 (14%) |
| Health Services Research | 15 (4%) | 2 (6%) |
| Medicine | 30 (8%) | 2 (6%) |
| Biostatistics | 14 (4%) | 2 (6%) |
| Other | 69 (18%) | 3 (8%) |

### Knowledge

Over two-thirds of the Twitter respondents reported that they had received formal training in causal graphs, mostly from graduate school courses. The most common causal graphs participants received training on were directed acyclic graphs [1], less than 20% received training on SWIGS [9,10] and an even smaller proportion of participants were trained on the use of FFRCISTGs [11].

Responses to the knowledge assessment questions were mixed (Table 2 and Table 3). Half of respondents correctly identified that not drawing an arrow between two variables was a stronger assumption than drawing an arrow, which is a critical assumption for correct DAG usage. Excluding an arrow between two variables removes any possibility of a causal relationship between the two, whereas the inclusion of an arrow indicates a causal relationship but does not make any assumption on the strength of that relation. In other words, including an arrow leaves the potential for a causal relationship to exist and excluding an arrow does not allow for any possible causal relationship between two variables. Respondents who reported having received training on causal graphs were more likely to choose the correct answer for this question and were less likely to be unsure about the answer than individuals who reported no training (Figure 1). Interestingly, while individuals who had completed an epidemiology program or who were current students in an epidemiology program were more likely to choose the correct answer than individuals in other fields, the type of training (graduate school course, workshop, or online training) did not appear to be associated with level of knowledge.

**Table 2.** Percentage of respondents from each survey version who correctly identified that not drawing an arrow on a DAG is a stronger assumption than drawing an arrow.

|  | Original Twitter Link<br>(%) | SER-disseminated Link<br>(%) |
| --- | --- | --- |
| **Student Status** |  |  |
| Current Student | 47.12 | 71.43 |
| Not a Student | 52.94 | 73.33 |
| **Degree Field** |  |  |
| Epidemiology | 56.02 | 75.00 |
| Other Field | 44.35 | 70.00 |
| **Training on DAGs** |  |  |
| Received Training | 57.69 | 70.83 |
| No Training | 34.57 | 85.71 |

**Table 3.** Correct responses to the question “Which of the following directed acyclic graphs, if correct, would imply that the true causal effect of post-traumatic stress disorder (PTSD) on suicide could be estimated without error? (select all that apply)” by student status, degree field and training. For both surveys, **the correct choice was DAG B (Figure 2b)**.

|  | Correct (Twitter Survey) | Correct (SER Link) |
| --- | --- | --- |
| **Student Status** |  |  |
| Current Student | 71.00% | 71.40% |
| Not a Student | 57.80% | 63.60% |
| **Degree Field** |  |  |
| Epidemiology | 60.90% | 66.70% |
| Other Field | 64.50% | 66.70% |
| **Training on DAGs** |  |  |
| Received Training | 62.20% | 75.00% |
| No Training | 60.90% | 66.70% |

**Figure 1:**
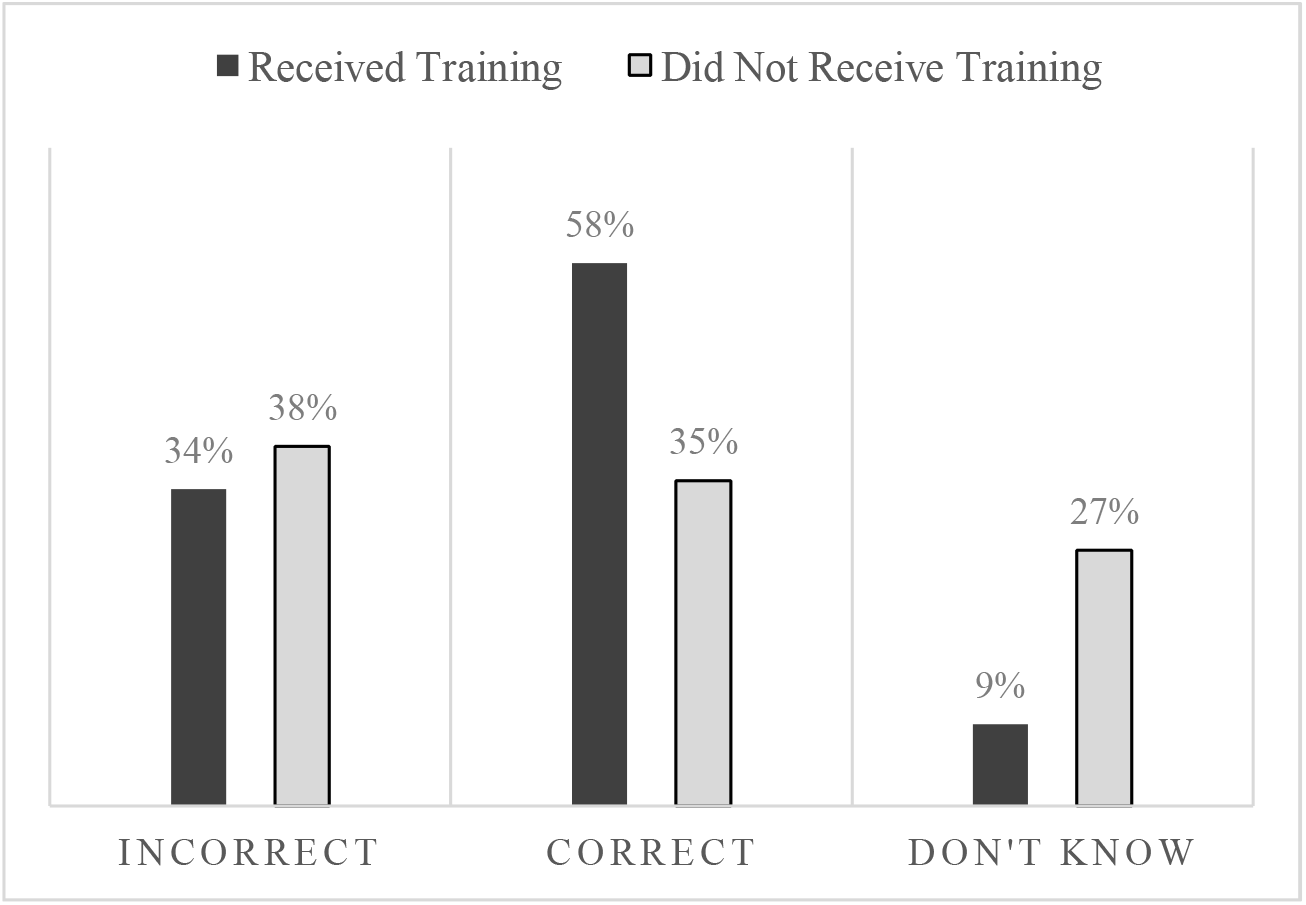
Bar graph of responses to the question “To the best of your understanding, which of the following is the stronger assumption in a directed acyclic graph (DAG)?” by causal graph training status. Possible responses were “Drawing an arrow between two variables” (incorrect); “Not drawing an arrow between two variables” (correct) and “I don’t know”.

A second knowledge question was designed to assess understanding of the depiction of measurement error in DAGs. When asked to choose which of two DAGs (Figure 2), if true, allowed identification of the causal effect of post-traumatic stress disorder on suicide, 62% of respondents chose the graph depicted in Figure 2b over that depicted in Figure 2a. This was the answer we had intended for respondents to select, because the DAG in Figure 2a represents intractable confounding by the true value of the confounder even when the measured confounder is included in the analysis or study design. However, subsequent to our study, it was brought to our attention that the DAG in Figure 2b requires strong assumptions about the strength of the direct effect of the measured exposure on the outcome in order to allow testing of the causal effect, and thus it would have also been reasonable to assume this DAG did not allow effect identification. Given the wording of the question, neither answer is exactly correct. Despite this, the majority of respondents did indeed select the DAG in Figure 2b as allowing effect identification, and there was essentially no difference in performance on this question between individuals who did and did not report having received training in causal graphs. However, current students were more likely to choose Fig 2b than non-students (Table 3). Further exploration of how well researchers (including ourselves) understand complex applied DAGs is warranted.

**Figure 2:**
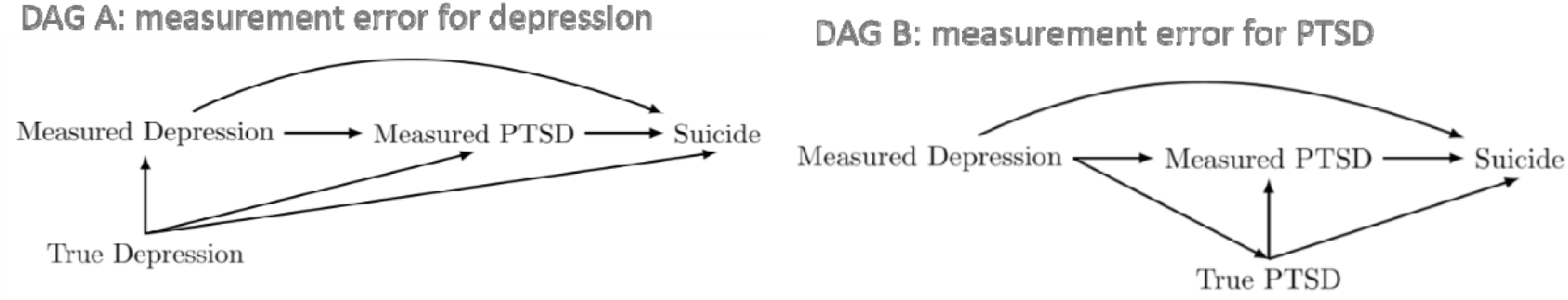
Suggested directed acyclic graphs (DAGs) demonstrating assumptions about measurement error for depression and post-traumatic stress disorder (PTSD). Survey respondents were asked “Which o the following directed acyclic graphs, if correct, would imply that the true causal effect of post-traumatic stress disorder (PTSD) on suicide could be estimated without error? (select all that apply)”. For response summary, see Table 2. **DAG B was intended as the correct response**.

### Attitudes

Most respondents somewhat or strongly agreed that they were comfortable using causal graphs for designing data collection, identifying appropriate adjustments sets, evaluating existing studies, and assessing surprising study results, regardless of training history. Nearly 80% of respondents somewhat or strongly agreed that causal graphs were useful in a classroom setting for describing bias, in an applied research project at the study design or analysis phase, or in reviewing a paper or critiquing an existing study. Over half of participants indicated that a more complicated DAG was best for designing a research study (Figure 3c) and least useful for presenting study assumptions. The simpler DAGs shown in Figure 3a and Figure 3b were rated as being more useful for presenting results.

**Figure 3:**
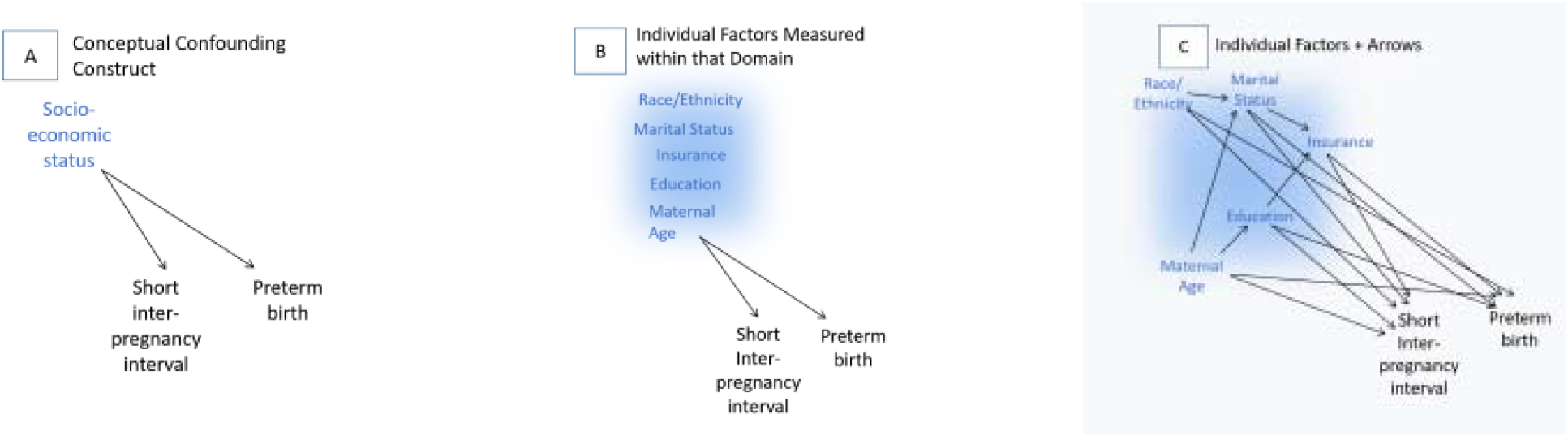
Three approaches to drawing directed acyclic graphs (DAGs) for study design, and presentation of research assumptions. Survey respondents were asked to rank the options in order from most (1) to least (3) useful for presenting research results and from most (1) to least (3) useful for designing the study.

Effect measure modification (EMM) and interaction are often of interest in epidemiology, but there are differing approaches to visualizing and assessing these features in a causal DAG, even amongst our research team [3]. A few of the arguments are that (1) arrows are agnostic to the presence or absence of EMM and interaction and therefore all arrows by definition represent the potential for EMM or interaction [1]; (2) EMM and interaction sometimes arise from the presence of alternate causes of the outcome of interest (that may or may not vary in distribution by exposure status), and so this type of EMM can be displayed by including unknown causes of the outcome [3,12]; and finally (3) EMM can be indicated by drawing an arrow from the modifier to another arrow, instead of to a node (note that these are no longer DAGs, and should be referred to more generally as ‘causal graphs’) [13]. When we asked respondents “to the best of your understanding, can effect modification be represented in a directed acyclic graph,” there was substantial disagreement (Table 4). We did not consider responses to this question to be correct or incorrect; we posed this question to get an understanding of the attitudes toward EMM representation on DAGs in our study population. In our experience, representation of EMM on DAGs is rarely taught. Despite this, individuals with any training were generally confident in their ability to choose one particular answer, and less likely to select “don’t know” compared to those with no training. However, the answers selected were nearly evenly split across the three choices, highlighting a potential lack of consensus in the field (Table 4).

**Table 4.**
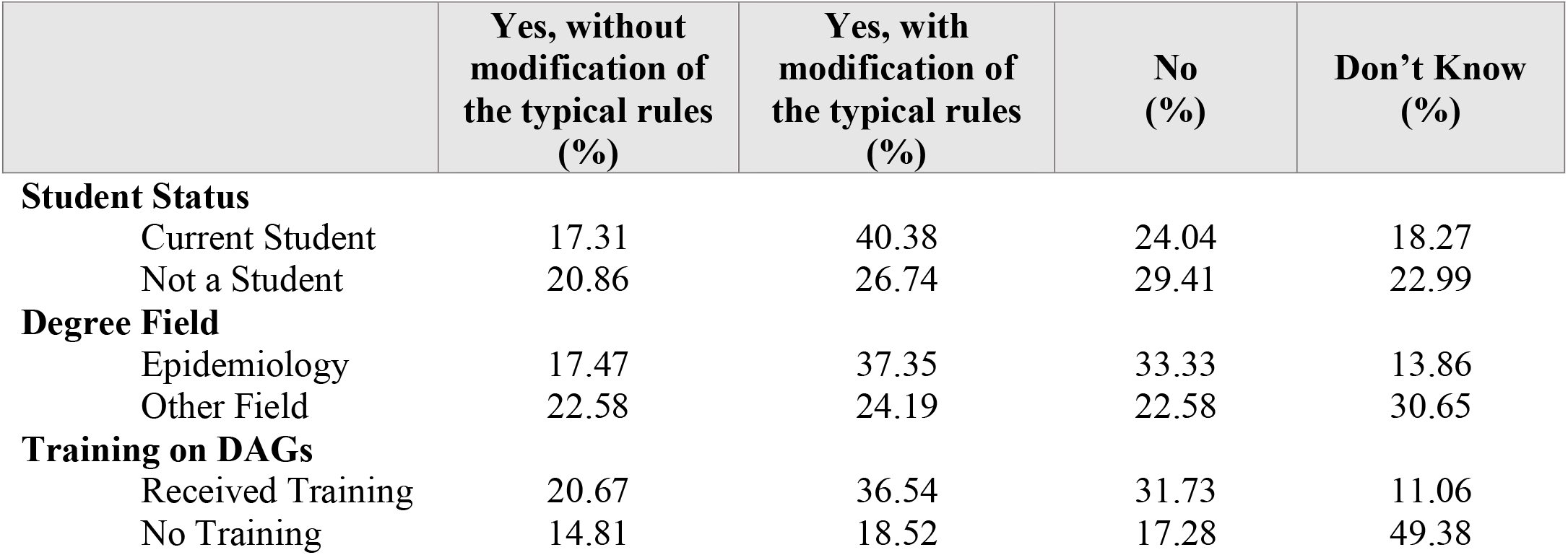
Responses to the question “To the best of your understanding, can effect modification be represented in a directed acyclic graph?” from the original Twitter survey by student status, degree field, and training type. There was no correct answer for this question.

|  | **Yes, without**<br>**modification of**<br>**the typical rules**<br>**(%)** | **Yes, with**<br>**modification of**<br>**the typical rules**<br>**(%)** | **No**<br>**(%)** | **Don’t Know**<br>**(%)** |
| --- | --- | --- | --- | --- |
| **Student Status** |  |  |  |  |
| Current Student | 17.31 | 40.38 | 24.04 | 18.27 |
| Not a Student | 20.86 | 26.74 | 29.41 | 22.99 |
| **Degree Field** |  |  |  |  |
| Epidemiology | 17.47 | 37.35 | 33.33 | 13.86 |
| Other Field | 22.58 | 24.19 | 22.58 | 30.65 |
| **Training on DAGs** |  |  |  |  |
| Received Training | 20.67 | 36.54 | 31.73 | 11.06 |
| No Training | 14.81 | 18.52 | 17.28 | 49.38 |

### Practices and barriers

About 60% of participants responded that they used causal graphs in an applied epidemiology project at least sometimes. Of these participants, only 20% responded that they used systematic processes (literature reviews or expert consensus) to develop a graphical causal model. The remaining 80% either performed informal literature reviews and conversations or re-used already published models.

Among the 40% of respondents that indicated they had never used a causal graph in an applied epidemiology project, the most common reason for not using a graphical causal model in applied epidemiology research was not knowing how to use them. When it came to challenges specifically associated with building causal graphs in epidemiology research, the most common challenge was choosing which arrows to omit, followed by choosing which arrows to include. In terms of assessing causal graphs in research, over half of participants found identifying potential unknown sources of error and identifying collider bias to be the two most challenging aspects.

Respondents who reported regularly using causal graphs (“sometimes”, “often”, or “always” responses) in their research most frequently said reference material, software tools, and availability of pre-published causal graphs would be of most use in helping them decide to use causal graphs more often. On the other hand, respondents who did not use causal graphs (“rarely”, “once”, or “never” responses) said online training would be most valuable, followed by in-person training and software tools. Approximately twice as many respondents who reported not regularly using causal graphs indicated that journal or grant agency requirements to use causal graphs would be useful for their uptake of these methods, compared to respondents who reported regularly using causal graphs.

### Qualitative analysis of text responses

Individuals who reported that they did not feel comfortable using causal graphs for designing data collection, identifying adjustment sets, evaluating studies or assessing surprising study results were given the opportunity to explain what aspects they were not comfortable with and what resources might help. A common theme among the responses was lack of knowledge contributing to discomfort using causal graphs. One participant wrote, “I haven’t received training in them at all but I feel like I should know more about them.” Other responses included more specific knowledge gaps. For example, one participant expressed concern about “How to incorporate effect modification, simultaneous bias, measurement error,” and another said, “Knowledge about how to correctly use the arrows.” Additionally, many participants wanted resources and training including a “resource library,” “access to experts,” and “online training materials.”

Participants were also asked to expand on the usefulness of causal graphs in classrooms, designing research, analyzing results, and reviewing completed work. Many respondents addressed a lack of knowledge about causal graphs, with some general statements such as “I should study it more” and “I do not understand how it would be used in this context.” Some participants mentioned more specific gaps in knowledge including “how to depict moderation.” Additionally, several respondents expressed concern about the methodological components of causal graphs including that “some forms of bias are not amenable to being described by DAGs” and “molecular biology is often bi-directional which is never represented.” There were also a few participants who expressed concern over using DAGs for analysis, stating “analyses have to be done based on model formulas, not on DAGs” and “at the study analysis phase you stick to the analysis plan.”

### Society for Epidemiologic Research survey results

In general, the responses from the Society for Epidemiologic Research-disseminated survey link were similar to those from the original Twitter survey. Most of the respondents in the Society for Epidemiologic Research-disseminated survey link reported using causal graphs in their research at least “sometimes”. Of these participants, almost all participants reported using informal methods (literature reviews and conversations) or re-using already published models to develop models for their research. Aligned with the original Twitter survey responses, reference materials were cited as the most useful tool to increase DAG usage among those who already use DAGs in research in the Society for Epidemiologic Research-disseminated survey. The most notable difference between the two surveys was that a higher proportion of respondents of the Society for Epidemiologic Research-disseminated survey link correctly answered that not drawing an arrow between two variables was a stronger assumption than drawing an arrow. Similar to the Twitter-distributed survey results, holding an epidemiology degree and being a student did not appear to be associated with correct responses (Table 4).

## Discussion

A large proportion of individuals surveyed who professionally engage in applied epidemiology agreed that causal graphs were useful for designing, analyzing and reading epidemiologic studies. Most of the respondents had been formally trained on causal graphs, the most common type being DAGs, and a majority expressed comfort in using causal graphs in their own research. However, only half of respondents correctly responded to the question on relational assumptions in DAGs. This suggests a gap between utilization of DAGs and knowledge about the proper use of DAGs. Those who received training appeared to have better knowledge about DAG operationalization, regardless of type of training, which may provide some support for developing additional trainings on causal graphs in a variety of formats, including online courses, reference books and in-person trainings. Further, most of the respondents who regularly use DAGs do not use a systematic process for developing the models, which may result in inaccurate DAGs and thus potentially fatal bias in studies based off those DAGs. The most common reason for not using DAGs in applied epidemiology projects was lack of knowledge. Those who did not use DAGs thought online or in-person trainings, as well as software tools, would help them use causal graphs more often.

There was disagreement about the representation of effect measure modification in DAGs, mirroring the debate within our research team. Identifying effect measure modification is often an important part of a causal research questions and is thus an important concept to understand when building causal graphs. The variety of attitudes on representing effect measure modification in our survey responses, paired with the finding that many participants reported re-using already published DAGs for their own research, illustrate the need for consensus within the epidemiology community [3]. We were not able to explore the attitudes towards effect measure modification in DAGs further and propose this as an area of future research.

Our findings build upon the findings of a recent, large, systematic review of published causal graphs in the epidemiologic literature [2]. There, the authors found that 38% of papers which claimed to have used DAGs did not include the model in the main text or supplementary material. Of the DAGs that were published, there were significant variations in terms of model design (i.e., number of nodes/arrows and use of unobserved variables) and DAG-based adjustment techniques. This may be explained, in part, by our finding that preference for the design of a research DAG varied depending on the purpose of the DAG (whether for designing a research study or for presenting study assumptions) (Figure 3). Additionally, the authors of the literature review reported that extraction errors were reduced when diagrams were constructed using DAGitty, a web-based platform for designing and exploring causal diagrams [14].

### Future directions

Our results demonstrate a need for more resources and guidance on the application of DAGs to epidemiology research. Although interest in and utilization of causal graphs is high, the lack of available training resources possibly contributes to the confusion and disagreement about DAG assumptions and rules. Promisingly, we saw relatively little difference in the level of knowledge about DAGs among those who had received graduate school training, workshop training, or online training, suggesting that increasing the availability of even a relatively low-intensity training may have substantial impact on the knowledge, and thus use of, causal graphs in epidemiology research. Finally, our results also suggest that improved guidance on the visualization and assessment of effect measure modification or interaction in causal graphs is necessary to address the uncertainty about this topic. Future research should also be focused on identifying explicit topics (i.e., expressing complex mixtures in DAGs) and training resources for researchers interested in using DAGs for study design and analysis.

### Limitations

One potential limitation of this study is that the sample is unlikely to be representative of all epidemiologists and medical researchers. In particular, it appears that women may have been somewhat more likely to participate than men, and current students may be over-represented. However, a study published in 2018 found that a higher proportion of epidemiology doctoral students, faculty members and society members were female and so this apparent imbalance may be representative of the field [15].The sample of participants who completed the survey may not be generalizable to the population of all practicing epidemiologists. The subset of epidemiologists who use Twitter may be younger than those who do not, and, if so, may be more likely to have been exposed to causal inference topics (including causal graphs) during their graduate training. However, we are aware of no previous research on the application of causal graphs among epidemiologists, and as such this study will be a valuable guide to the development of more systematic assessments and the creation of tools to improve the implementation of causal graphs in applied epidemiologic research. Additionally, our questions were not externally validated to measure DAG knowledge, and while the questions asked in this survey were based off of years of teaching experience on DAGs and participants were able to expand on their responses via free form text, we may have misclassified DAG knowledge. Finally, our questions were created by researchers in the United States, and while we had participants from outside the United States complete this survey, the questions and possible responses were framed within the context of causal graph teaching practices in the United States. It is possible that these questions may not represent DAG teaching attitudes and practices in other countries.

## Conclusion

Overall, there is general agreement within the epidemiology community that graphic causal models are useful for both applied research and teaching purposes; however, a lack of training resources presents a barrier for their widespread use in practice. Generally, epidemiologists and medical researchers who received training in causal graphs were shown to have better knowledge about the underlying assumptions for building DAGs compared to those without any training, regardless of the intensity of training. Easily accessible resources that provide guidelines for creating evidenced-based DAGs are likely needed to increase DAG utilization, which may improve analytic adjustment for confounders in published epidemiologic research.

## Data Availability

The data will not be publicly available under the terms of the informed consent given by survey respondents.

## Acknowledgements

The authors would like to thank the survey participants, the Society for Epidemiological Research, and everyone on #epitwitter who helped spread the word about our survey.

