## Supplementary material for "Assessing Knowledge, Attitudes, and Practices towards Causal Directed Acyclic Graphs: a qualitative research project": Online Supplemental Material

**Appendix 1: Recruitment Tweets**

Tweet on August 7^th^, 2019: “Hello #epitwitter #statstwitter & #medtwitter! Do you do applied epi or epi-related research? We're doing a research study on causal graphs & need your help! Take our survey: <https://bostonu.qualtrics.com/jfe/form/SV_6AsKw3bkkx3l21D>

Or have questions? Ask below!”

Tweet on August 8^th^, 2019: “We've already gotten 75 responses to our research survey and the results are starting to look super interesting, but we want more!! If you do epi/epi-related research, have ~10 min to spare, and haven't already participated, please check it out!”

Tweet on August 10^th^, 2019: “Do you do research on the health of populations? Access to medical services? Best treatment options? Prevalence of disease? Etc? Have you taken our research survey yet? It’s <10min & it will help us help you make science better!!”

Tweet on August 12^th^, 2019: “We've got 171 responses to our research survey! Can we get to 200?! If you haven't already, check it out here.”

Tweet from @societyforepi on November 20^th^, 2019 (same text and date as the Facebook post): Do you participate in applied medical or public health research? If so, please consider filling out this short survey on the use of graphical causal models, such as directed acyclic graphs, for research purposes. More info can be found through this link: https://bostonu.qualtrics.com/jfe/form/SV_8iBthMISjYgrg0J

**Appendix 2: Original Twitter Survey items**

*Introduction* Thank you for participating in our research study. We are hoping to explore issues related to the use of graphical causal models, such as directed acyclic graphs (DAGs) in applied epidemiologic research. The information we collect from this interview will be used in publications and to inform the development of tools and resources to improve the use of graphical causal models in epidemiology. By continuing the survey, you acknowledge your consent to participate in this research study. We will not collect IP addresses or other identifying information. The results of the survey may be published. Any published quotes will be anonymized to prevent identification.   For more information on the study and to read the consent form, please click [here](https://bostonu.qualtrics.com/CP/File.php?F=F_1KOpcwzWUF4V9CB).

Q1  Please click the circle to indicate that you consent to participate in this survey. If you do not consent, please exit the survey now.

- I Consent

Condition: **I Consent** **Is Not Selected**. Skip To: **Thank you. To submit your answers, pl...**.

*Part 1: Background and Research History* We would like to collect some optional information about you, your background, and your research history. We will use this information to help understand the study population, and to identify groups that may benefit from targeted tool, guidance, or training material development.

Q2 Which best describes your current gender?

- Man (1)
- Woman (2)
- Indigenous or other cultural gender minority identity (e.g. two-spirit) (3)
- Non-binary (4)
- Gender-fluid (5)
- Other [please describe] (6) ________________________________________________
- Prefer not to say (7)

Q3 Please select any of the following racial / ethnic identities as appropriate: (Select all that apply)

- Black, including African American and African (1)
- Asian, including East Asian and South Asian (2)
- Middle Eastern or Northern African (3)
- Pacific Islander or Native Hawaiian (4)
- Indigenous, including Native American, First Nations, Aboriginal, Metis (5)
- Hispanic (6)
- White (7)
- Other [please describe] (8) ________________________________________________
- Prefer not to say (9)

Q4 Are you currently a student?

- Yes (1)
- No (2)

Display This Question:

If Are you currently a student? = Yes

Q5 Which program are you currently enrolled in?

- Bachelors (1)
- Masters of Science (2)
- Masters of Public Health (3)
- Masters of Arts (4)
- PhD or ScD/ DSc or equivalent (5)
- Medical doctorate (6)
- Combined MD-PhD or equivalent (7)
- Other [please describe] (8) ________________________________________________
- Prefer not to say (9)

Display This Question:

If Are you currently a student? = Yes

Q6 What field of study is this program in?

- Epidemiology (1)
- Public Health (2)
- Medicine (3)
- Health Services Research (4)
- Biostatistics (5)
- Other [please describe] (6) ________________________________________________
- Prefer not to say (7)

Display This Question:

If Are you currently a student? = No

Q7 What is your highest level of education?

- Bachelors (1)
- Masters of Science (2)
- Masters of Public Health (3)
- Masters of Arts (4)
- PhD or ScD/ DSc or equivalent (5)
- Medical Doctorate (6)
- Combined MD-PhD or equivalent (7)
- Other [please describe] (8) ________________________________________________
- Prefer not to say (9)

Display This Question:

If Are you currently a student? = No

Q8 What field is your highest degree in?

- Epidemiology (1)
- Public Health (2)
- Medicine (3)
- Health Services Research (4)
- Biostatistics (5)
- Other [please describe] (6) ________________________________________________
- Prefer not to say (7)

Q9 What country do you primarily work, practice or study epidemiology in currently?

- United States of America (1)
- Canada (2)
- Other [please describe] (3) ________________________________________________
- Prefer not to say (4)

Q10 How would you describe your current position?

- Academia (1)
- Government – local (2)
- Government – state / provincial / regional (3)
- Government – federal /national (4)
- Government – international (5)
- Non-Governmental Organization (NGO) (6)
- Pharmaceutical company (7)
- Consulting or Contract Research Organization (CRO) (8)
- Other [please describe] (9) ________________________________________________
- Prefer not to say (10)

*Part 2: Knowledge.* The next set of questions are designed to assess your current level of knowledge about graphical causal models.

Q11 Did you ever receive formal training on the use of any graphical causal model?

- Yes, in graduate school (1)
- Yes, at a workshop, conference, or similar (2)
- Yes, from an online training (3)
- No (4)
- Other [please describe] (5) ________________________________________________

Q12 Did you ever receive training on the use of directed acyclic graphs (DAGs)?

- Yes, in graduate school (1)
- Yes, at a workshop, conference, or similar (2)
- Yes, from an online training (3)
- No (4)
- Other [please describe] (5) ________________________________________________

Q13 Did you ever receive training on the use of finest fully randomized causally interpretable structured tree graphs (FFRCISTGs)

- Yes, in graduate school (1)
- Yes, at a workshop, conference, or similar (2)
- Yes, from an online training (3)
- No (4)
- Other [please describe] (5) ________________________________________________

Q14 Did you ever receive formal training on the use of single world intervention graphs (SWIGs)

- Yes, in graduate school (1)
- Yes, at a workshop, conference, or similar (2)
- Yes, from an online training (3)
- No (4)
- Other [please describe] (5) ________________________________________________

Q15 To the best of your understanding, which of the following is the stronger assumption in a directed acyclic graph (DAG)?

- Drawing an arrow between two variables (1)
- Not drawing an arrow between two variables (2)
- Don’t know (3)

Q16 To the best of your understanding, can effect modification be represented in a directed acyclic graph?

- Yes, without modification of the typical rules (1)
- Yes, with modification of the typical rules (2)
- No (3)
- Don’t know (4)

Q17
Rank the following directed acyclic graphs from most (1) to least (3) useful *for designing* a research study (drag and drop the graphs into your preferred order with 1 at the top)

______
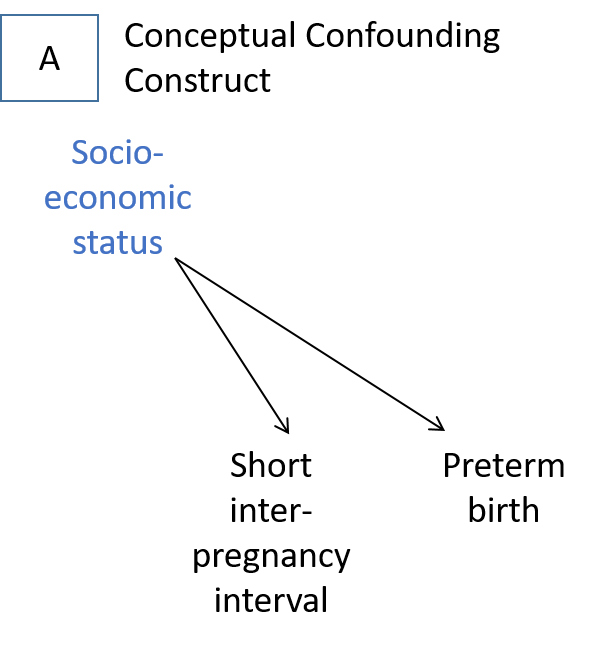

______
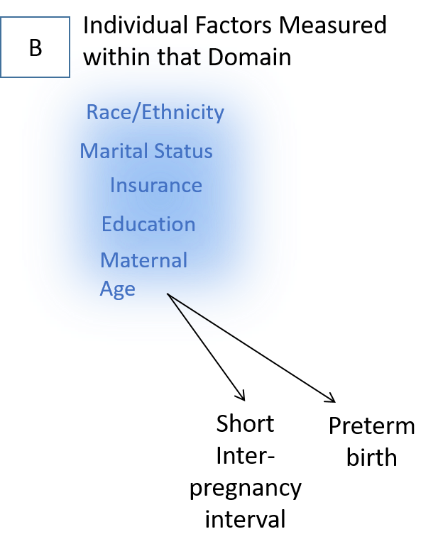

______
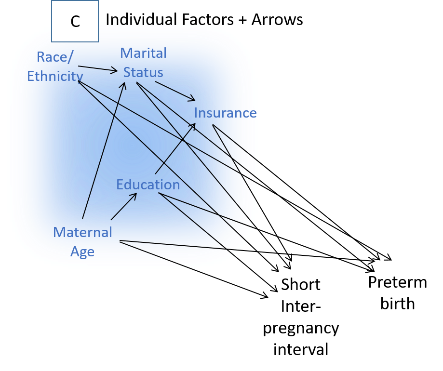

Q18
Rank the following directed acyclic graphs from most (1) to least (3) useful for *presenting the results* of a research study (drag and drop the graphs into your preferred order with 1 at the top)

______
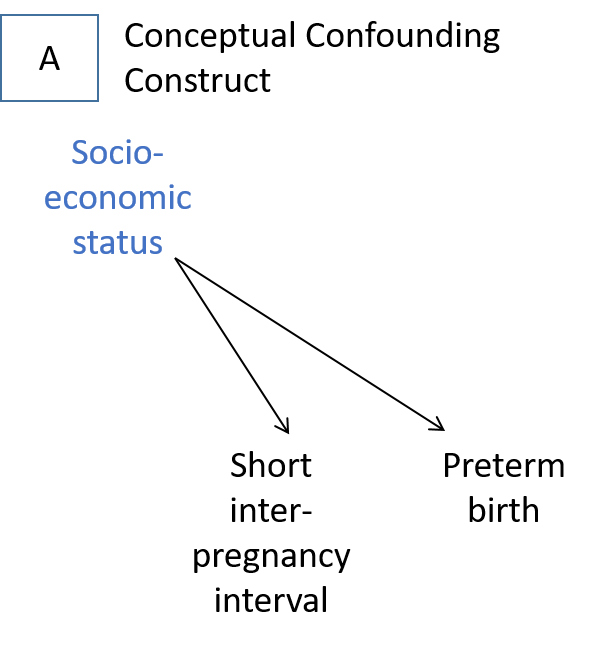

______
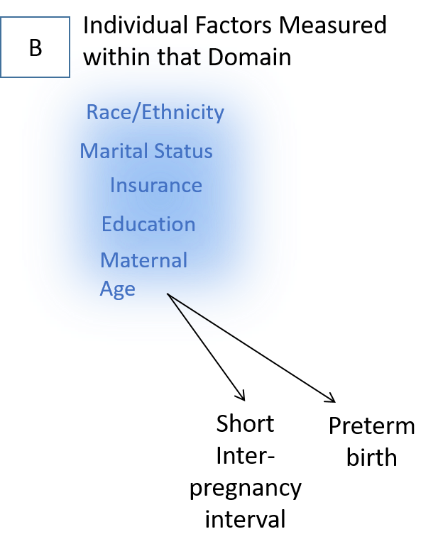

______
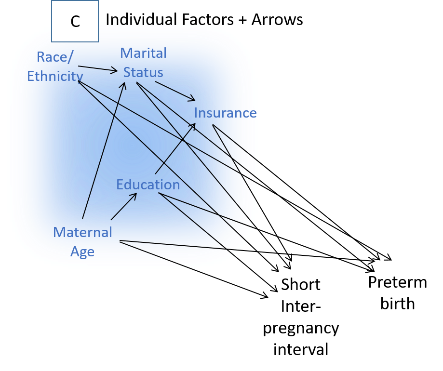

Q19 Which of the following directed acyclic graphs, if correct, would imply that the true causal effect of post-traumatic stress disorder (PTSD) on suicide could be estimated without error? (select all that apply)

-
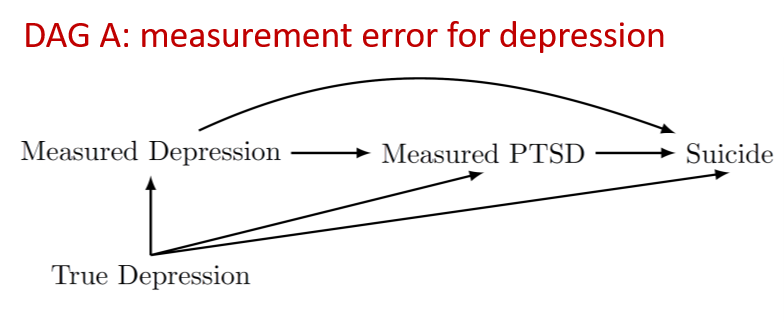

-
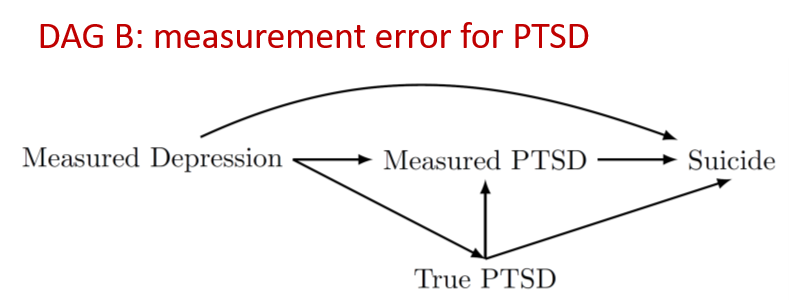

Q20 *Part 3: Attitudes.* The following questions are designed to assess your current attitudes towards graphical causal models. Please select the appropriate level of agreement with each statement below.

Q21 I am comfortable using graphical causal models .... (if you are viewing on mobile, please click to expand each item below)

|  | Strongly agree (1) | Somewhat agree (2) | Neither agree nor disagree (3) | Somewhat disagree (4) | Strongly disagree (5) |
| --- | --- | --- | --- | --- | --- |
| ...for designing data collection (1) |  |  |  |  |  |
| ...for identifying appropriate adjustment sets (2) |  |  |  |  |  |
| .... for evaluating existing studies (3) |  |  |  |  |  |
| ...for assessing surprising study results (4) |  |  |  |  |  |

Q22 If you answered "somewhat disagree" or "strongly disagree" to any of the prompts in the above question, what aspect are you not comfortable with and what resources would you like to have access to for increasing your comfort level?

Q23 I find graphical causal models useful ... (if you are viewing on mobile, please click to expand each item below)

|  | Strongly agree (1) | Agree (2) | Somewhat agree (3) | Neither agree nor disagree (4) | Somewhat disagree (5) | Disagree (6) | Strongly disagree (7) |
| --- | --- | --- | --- | --- | --- | --- | --- |
| ... in a classroom setting for describing biases (1) |  |  |  |  |  |  |  |
| ... in an applied research project at the study design phase (2) |  |  |  |  |  |  |  |
| ... in an applied research project at the study analysis phase (3) |  |  |  |  |  |  |  |
| ... when reviewing a paper or critiquing an existing study (4) |  |  |  |  |  |  |  |

Q24 If you answered "somewhat disagree" or "strongly disagree" to any of the prompts in the above question, what aspect do you find to not be useful and how could these be improved or addressed?

________________________________________________________________

**Part 4: Practices.** The final set of questions are designed to assess your current practices regarding graphical causal models

Q25 Have you ever used a graphical causal model in an *applied* epidemiology project (whether or not that project has resulted in a published manuscript)?

- Always (1)
- Often (2)
- Sometimes (3)
- Rarely (4)
- Once (5)
- Never (6)

Display This Question:

If Have you ever used a graphical causal model in an applied epidemiology project (whether or not th... = Rarely

Or Have you ever used a graphical causal model in an applied epidemiology project (whether or not th... = Once

Or Have you ever used a graphical causal model in an applied epidemiology project (whether or not th... = Never

Q26 Which of the best describes why you do not use graphical causal models in your applied epidemiology research? (Select all that apply)

- I don't like them (1)
- I don't know how to use them (2)
- I don't feel that I need them (3)
- They take too long to develop (4)
- My collaborators don't want to use them (5)
- Don't do applied epidemiologic research (6)
- Other [please describe] (7) ________________________________________________

Display This Question:

If Have you ever used a graphical causal model in an applied epidemiology project (whether or not th... = Rarely

Or Have you ever used a graphical causal model in an applied epidemiology project (whether or not th... = Once

Or Have you ever used a graphical causal model in an applied epidemiology project (whether or not th... = Never

Q27 Which of the following would help you decide to use graphical causal models more often? (Select all that apply)

- In-person training (1)
- Online training (2)
- Reference material (book, paper) (3)
- Software tools (4)
- Information to share with collaborators (5)
- Availability of consensus or pre-published graphical causal models (6)
- Journal or grant agency requirement (7)
- Nothing (I will not use them more) (8)
- Other [please describe] (9) ________________________________________________

Display This Question:

If Have you ever used a graphical causal model in an applied epidemiology project (whether or not th... = Always

Or Have you ever used a graphical causal model in an applied epidemiology project (whether or not th... = Often

Or Have you ever used a graphical causal model in an applied epidemiology project (whether or not th... = Sometimes

Q28 What are the primary reasons that you use graphical causal models in your applied epidemiologic research? (Select all that apply)

- Useful for designing data collection (1)
- Useful for designing analyses (2)
- Useful for understanding surprising results (3)
- My collaborators require it (4)
- Other [please describe] (5) ________________________________________________

Q29 Please rank the following from 1 (most challenging) to 5/6 (least challenging) in terms of the difficulty for building graphical causal models in your research
(drag and drop into your preferred order with 1 at the top)
______ Choosing variables (1)

______ Ordering variables (2)

______ Choosing which arrows to include (3)

______ Choosing which arrows to omit (4)

______ Defining variables (5)

______ Other [please describe] (6)

Q30 Please rank the following from 1 (most challenging) to 5 (least challenging) in terms of the difficulty for assessing graphical causal models *in your research*
(drag and drop into your preferred order with 1 at the top)
______ Identifying confounding paths (1)

______ Identifying collider bias (2)

______ Identifying analytic sets (3)

______ Identifying potential unknown sources of error (4)

______ Understanding variable meanings (5)

______ Other [please describe] (6)

Q31 Please rank the following from 1 (most challenging) to 5 (least challenging) in terms of the difficulty for assessing graphical causal models in *other people’s research* (drag and drop into your preferred order

with 1 at the top)
______ Identifying confounding paths (1)

______ Identifying collider bias (2)

______ Identifying analytic sets (3)

______ Identifying potential unknown sources of error (4)

______ Understanding variable meanings (5)

______ Other [please describe] (6)

Q32 What process do you currently use to develop a graphical causal model for your research? (Select all that apply)

- Systematic literature review (1)
- Systematic expert consensus process (2)
- Informal literature review (3)
- Informal discussion with experts (4)
- Re-use own published models (5)
- Re-use other people’s published models (6)
- Other [please describe] (7) ________________________________________________

Q33 Do you use any software to help you develop or assess graphical causal models? (Select all that apply)

- Yes, DAGgity (1)
- Yes, ggDAG (2)
- Yes, ShinyDAG (3)
- Yes, other [please describe] (4) ________________________________________________
- None or only for digital creation (e.g. Word, LaTeX) (5)

Q34 Which of the following would help you decide to use graphical causal models more often? (Select all that apply)

- In-person training (1)
- Online training (2)
- Reference material (book, paper) (3)
- Software tools (4)
- Information to share with collaborators (5)
- Availability of consensus or pre-published graphical causal models (6)
- Journal or grant agency requirement (7)
- Nothing (I already always use them) (8)
- Other [please describe] (9) ________________________________________________

Thank you. To submit your answers, please click 'Next'. If you would like to change any of your answers, you may do so by using the 'Back' button before you submit.

**Appendix 3: Additional items that were added to the Society for Epidemiologic Research-disseminated survey that were not in the original version**

Q35 Where did you hear about this survey?

- Society of Epidemiologic Research (SER) (1)
- American Public Health Association (APHA) (2)
- American College of Epidemiology (ACE) (3)
- EpiMonitor (4)
- Other (5) ________________________________________________
- Prefer not to say (6)

Q36 From the DAG below, which of the sets of variables below would control for confounding (select all that apply):

-
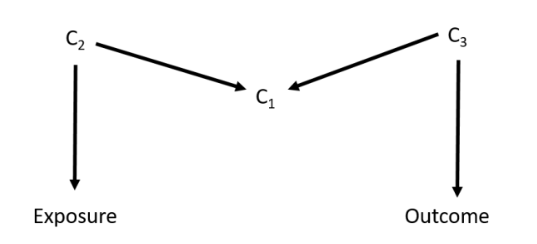
The null set (1)
- C1 (2)
- C1 and C2 (3)
- C1, C2 and C3 (4)
- C2 and C3 (5)

Q37 From the DAG below, which variable(s) should you adjust for to remove confounding (select all that apply):

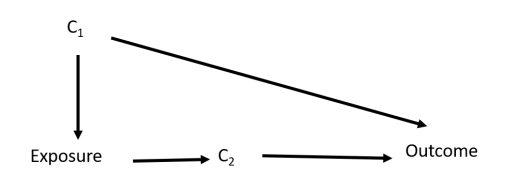

- C1 (1)
- C2 (2)
- C1 and C2 (3)
- None (4)

Q38 In which one of the following DAGs does the exposure have an identifiable effect on the outcome (a box indicates conditioning on that variable):

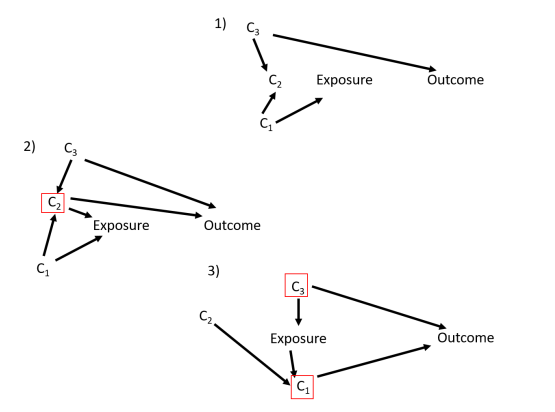

- 1 (1)
- 2 (2)
- 3 (3)
- 1 and 2 (4)
- 2 and 3 (5)
- 1 and 3 (6)
- 1, 2 and 3 (7)
